# Amplification of human β-glucoronidase gene for appraising the accuracy of negative SARS-CoV-2 RT-PCR results in upper respiratory tract specimens

**DOI:** 10.1101/2020.05.20.20105312

**Authors:** Eliseo Albert, Blanca Ferrer, Ignacio Torres, Alicia Serrano, María Jesús Alcaraz, Javier Buesa, Carlos Solano, Javier Colomina, Felipe Bueno, Dixie Huntley, Beatriz Olea, Arantxa Valdivia, David Navarro

## Abstract

Real-time reverse transcription polymerase-chain reaction (RT-PCR) is the mainstay of Covid-19 diagnosis. False-negative RT-PCR results may hamper clinical management of patients and hinder the adoption of epidemiological measures to control the pandemic. The current study was aimed at assessing whether amplification of β-glucoronidase (GUSB) gene would help estimate the accuracy of SARS-CoV-2 RT-PCR negative results in upper respiratory tract (URT) specimens. URT specimens that tested negative by SARS-CoV-2 RT-PCR displayed higher GUSB RT-PCR cycle thresholds (CT) (P=0.070) than those testing positive (median, 30.7; range, 27.0-40.0, and median 29.7; range 25.5-36.8, respectively), this reflecting poorer cellularity. Receiver operating characteristic (roc) curve analysis indicated that a CT threshold of 31.2 discriminated best between positive and negative SARS CoV-2 RT-PCRs (area under a curve, 0.66; 95% CI, 0.50-0.81; *P*=0.08). This cut-off yielded a true negative ratio of 89% and accuracy of 70%. The data suggested that amplification of the GUSB gene by RT-PCR may help to appraise the accuracy of negative SARS-CoV-2 RT-PCR results in patients in whom Covid-19 is eventually diagnosed.

---

Real-time reverse transcription polymerase-chain reaction (RT-PCR) is the mainstay of Covid-19 diagnosis.^1^ Up to 30% of patients clinically suspected of Covid-19 may have initial or repeat RT-PCR negative results prior to positive test conversion, most notably when upper respiratory tract (URT) specimens are processed.^2-7^ False-negative RT-PCR results may hamper clinical management of patients and hinder the adoption of epidemiological measures to control the pandemic. A number of pre-analytical and analytical factors may impact on the diagnostic efficiency of RT-PCR, including the type of and time to specimen processing, conservation prior to testing, quality of samples, timing of sample collection after symptoms onset or the intrinsic performance of the assay (i.e. limit of detection-LOD-).^2,8^ A large number of commercially-available SARS-CoV-2 RT-PCR assays targeting one or more SARS-CoV-2 genes have been launched and have reached global widespread use.^9^ Most of these assays include a spike-in control for RT-PCR amplification, such as MS2 phage RNA genome, but provide no information on specimen cellularity, since no primers targeting human housekeeping genes (i.e. RNAse P) are included in the reaction. The current study was aimed at assessing whether amplification of β-glucoronidase (GUSB) gene would help estimate the accuracy of SARS-CoV-2 RT-PCR negative results in URT samples.

As shown in Figure 1, 202 patients eventually diagnosed with Covid-19, either by RT-PCR (n=191) or serological methods (n=11), were admitted to our center (median age, 65 years; range, 3-98 years; 115 male and 87 female) until April 14. A total of 199 patients received a final diagnosis of Covid-19 on clinical, laboratory and imaging grounds, without microbiological documentation. The study was approved by the local Ethics Committee (INCLIVA).

**FIGURE 1.**
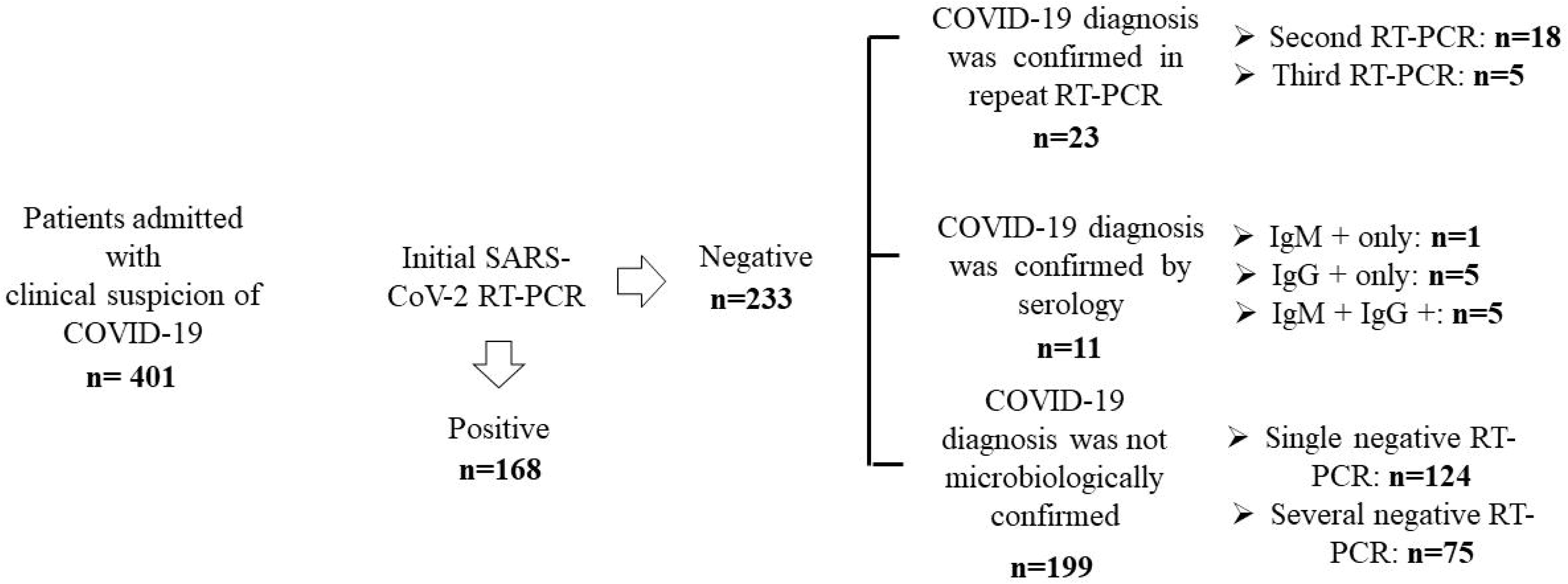
SARS-CoV-2 RT-PCR and serology testing results in patients with clinical suspicion of Covid-19 admitted to Hospital Clínico Universitario of Valencia during the study period. Nasopharyngeal or Oropharyngeal swabs were obtained with flocked swabs in universal transport medium (Beckton Dickinson, Sparks, MD, USA, or Copan Diagnostics, Murrieta, CA, USA) and conserved at 4 ° C until processed (within 6 hours). Nucleic acid extraction was performed using the Qiagen EZ-1 Viral extraction kit or the DSP virus Pathogen Minikit on the EZ1 or QiaSymphony Robot instruments (Qiagen, Valencia, CA, USA), respectively. Commercially-available PCR assays used for SARS CoV-19 testing included the LightMix® Modular SARS-CoV-2 (COVID-19) E-gene/LightMix® Modular SARS-CoV-2 (COVID-19) RdRP gene from TIB MOLBIOL GmHD, distributed by Roche Diagnostics (Pleasanton, CA, USA) on the Light Cycler 2.0 instrument, the SARS-COV-2 REALTIME PCR KIT from Vircell Diagnostics (Granada, Spain), or the REALQUALITY RQ-2019-nCoV from AB ANALITICA (Padua, Italy), both on the Applied Biosystems 7500 instrument and the SARS-CoV-2 (S gene) – BD MAX™ System (VIASURE Real Time PCR Detection Kits; CerTest, Zaragoza, Spain). Results were interpreted according to the respective manufacturer’s instructions.

Of the 202 patients, 34 (16.8%) tested negative by RT-PCR on first URT specimens, collected at a median of 5 days (range, 1-14 days) after the onset of symptoms. In these patients, as per protocol, URT swabs were collected every 24-72 h until RT-PCR positive conversion. Twenty-three patients tested positive by RT-PCR in the second (n=18) or third (n=5) URT sample. Diagnosis of Covid-19 was achieved by serological methods in the remaining 11 patients. Sera from these patients were drawn at a median of 12 days (range, 10-21 days) after admission. The presence of either SARS-CoV-2 IgM (determined by the MAGLUMI 2019-nCoV SARS-CoV-2-IgM assay on the fully automated MAGLUMI analyzers-SNIBE – Shenzhen New Industries Biomedical Engineering Co., Ltd, Shenzhen, China), IgG (Euroimmun anti-SARS-CoV-2 IgG assay; Euroimmun, Luebeck, Germany), or both confirmed diagnosis of Covid-19.

A total of 47 URT specimens (26 and 21 specimens yielding negative or positive SARS-CoV-2 RT-PCR results, respectively) from 21 of the above-mentioned 23 patients were subjected to GUSB gene RT-PCR analysis, which was performed in a parallel to viral RT-PCR testing. To this end, we used the HEQC-one step kit (Seqplexing, Valencia, Spain), a one-Step real-time RT-PCR. RNA was extracted from clinical samples using the DSP virus Pathogen Minikit on the QiaSymphony Robot instruments (Qiagen, Valencia, CA, USA), reverse transcribed to cDNA and subsequently amplified in the LightCycler 480 Real-Time PCR System Version II (Roche Diagnostics, Pleasanton, USA). Cy5 fluorescent signal (618-660 nm) revealed amplification of the target gene.

URT specimens that tested negative by SARS-CoV-2 RT-PCR displayed higher GUSB RT-PCR cycle thresholds (CT) (*P*=0.070; Mann-Whitney U test) than those testing positive (median, 30.7; range, 27.0-40.0, and median 29.7; range 25.5-36.8, respectively), this reflecting poorer cellularity. Receiver operating characteristic (roc) curve analysis (not shown) indicated that a C_T_ threshold of 31.2 discriminated best between positive and negative SARS CoV-2 RT-PCRs (area under a curve, 0.66; 95% CI, 0.50-0.81; *P*=0.08). This cut-off yielded a true negative ratio of 89% and accuracy of 70% (Table 1).

**TABLE 1.** Accuracy of negative SARS-CoV-2 RT-PCR results in upper respiratory tract specimens from patients with microbiological diagnosis of Covid-19 upon β-glucoronidase gene RT-PCR C_T_ value.

| β-glucoronidase gene $C_T$ | SARS-CoV-2 RT-PCR | |
| --- | --- | --- |
|  | Positive<sup>a</sup> | Negative<sup>b</sup> |
| $\leq 31.2$ | 18 | 18 |
| $> 31.2$ | 3 | 8 |

$C_T$ , RT-PCR cycle threshold.
<sup>a</sup>The REALQUALITY RQ-2019-nCoV was used in 7 specimens. The LightMix® Modular SARS-CoV-2 (COVID-19) E-gene/LightMix® Modular SARS-CoV-2 (COVID-19) RdRP gene was used in 6 specimens. The SARS-COV-2 REALTIME PCR KIT was used in 4 specimens. The SARS-CoV-2 (S gene) – BD MAX™ System (VIASURE Real Time PCR Detection Kits) was used in 4 specimens.
<sup>b</sup>The LightMix® Modular SARS-CoV-2 (COVID-19) was used in 17 specimens. The REALQUALITY RQ-2019-nCoV was used in 6 specimens. The SARS-COV-2 REALTIME PCR KIT was used in 3 specimens.

The current study has several limitations that should be acknowledged. First, a relatively scarce number of specimens were subjected to GUSB gene analysis. Second, consecutive specimens from a given patient could have been tested by different SARS-CoV-2 RT-PCR assays displaying distinct LOD. Third, regardless of the specimen cellularity, SARS CoV-2 load may have been intrinsically lower in patients testing negative than in those testing positive, although this possibility seems unlikely given the dynamics of viral load in URT specimens during the course of Covid-19, which peaks within the first week after onset of symptoms.^10,11^ In this sense, as stated, first and second SARS-CoV-2 RT-PCR negative specimens were usually collected within this time frame. In summary, our data suggested that amplification of the GUSB gene by RT-PCR may help to appraise the accuracy of negative SARS-CoV-2 RT-PCR results in patients in whom Covid-19 is eventually diagnosed. Further studies are warranted to validate this assumption.

## Data Availability

The data that support the findings of this study are avaliable from the corresponding author (DN) upon reasonable request

## ACKNOWLEDGMENTS

No public or private funds were used for the current study. We are grateful to all personnel working at Microbiology Service of Clinic University Hospital for their unwavering commitment in the fight against Covid-19. We are indebted to all colleagues who attended the patients, in particular to Josep Redón, María Luisa Blasco, Jaime Signes-Costa, María José Galindo, and María José Forner.

## AUTHOR CONTRIBUTIONS

EA, BF, IT, AS, and CS performed the GUSB gene RT-PCR analyses and designed the study. MJA performed the serological tests. JB, JC, FB, DH, BO, AV performed SARS-CoV-2 RT-PCR tests. DN designed the study, analyzed the data and wrote the manuscript. All authors reviewed and approved the final version of the manuscript.

## REFERENCES

1. Laboratory testing strategy recommendations for COVID-19: interim guidance. WHO/2019-nCoV/lab_testing/2020.1

2. Arevalo-Rodriguez I, Buitrago-Garcia D, Simancas-Racines D, et al. False-negative results of initial RT-PCR assays for Covid-19: a systematic review. *MedRxiv preprint* 2020; doi: https://doi.org/10.1101/2020.04.16.20066787.

3. Ai T, Yang Z, Hou H, et al. Correlation of Chest CT and RT-PCR Testing 448 in Coronavirus Disease 2019 (COVID-19) in China: A Report of 1014 Cases. Radiology. 2020: 200642.

4. Bernheim A, Mei X, Huang M, et al. Chest CT Findings in 451 Coronavirus Disease-19 (COVID-19): Relationship to Duration of Infection. Radiology. 2020: 200463.

5. Fang Y, Zhang H, Xie J, et al. Sensitivity of Chest CT for COVID-19: Comparison to RT-PCR. Radiology. 2020: 200432.

6. Li Y, Yao L, Li J, Chen L, Song Y, Cai Z, Yang C. Stability issues of RT-PCR testing of SARS-CoV-2 for hospitalized patients clinically diagnosed with COVID-19. J Med Virol. 2020; Mar 26. doi: 10.1002/jmv.25786.

7. Winichakoon P, Chaiwarith R, Liwsrisakun C, et al. Negative Nasopharyngeal and Oropharyngeal Swabs Do Not Rule Out COVID-19. J Clin Microbiol. 2020;58(5). pii: e00297–20.

8. Lowe CF, Matic N, Ritchie G, et al. Detection of low levels of SARS-CoV-2 RNA from nasopharyngeal swabs using three commercial molecular assays. J Clin Virol. 2020 Apr 28;128:104387.

9. Find Evaluation Update: SARS-COV-2 Molecular Diagnostics. https://www.finddx.org/covid-19/sarscov2-eval-molecular/.

10. Liu Y, Yan LM, Wan L, et al. Viral dynamics in mild and severe cases of COVID-19. Lancet Infect Dis. 2020; pii: S1473-3099(20)30232-2.

11. Zou L, Ruan F, Huang M, et al. SARS-CoV-2 Viral Load in Upper Respiratory Specimens of Infected Patients. N Engl J Med. 2020; 382: 1177-1179.

